# COVID-19 Reinfections in Mexico City: Implications for public health response

**DOI:** 10.1101/2022.12.08.22283269

**Authors:** Guillermo de Anda-Jauregui, Laura Gómez-Romero, Alberto Cedro-Tanda, Sofía Cañas, Abraham Campos-Romero, Jonathan Alcantar-Fernández, Alfredo Hidalgo-Miranda, Luis A. Herrera

**Author notes:** Correspondence (A.H.-M.); (L.A.H.); (A.H.-M.); (L.A.H.). These authors contributed equally to this work (Guillermo de Anda-Jauregui, Laura Gómez-Romero, Alberto Cedro-Tanda). The order of the authors was decided in consensus by the three first authors, but this order may be reported differently if it pleases them.

## Abstract

**Background:** SARS-CoV-2 pandemia continues to be important even when more than 60% of the global population has been vaccinated. As the pandemia evolves the number of reinfection cases will continue to increase as new variants are generated that evade the immune response. Understanding reinfections is important to guide the public health system and to inform decision-makers.

**Methods:** We downloaded clinical outcome and severity of infection data from the SISVER (respiratory disease epidemiological surveillance system) database. We sequenced SARS-CoV-2 samples, identified SARS-CoV-2 lineage and upload this genomic data to GISAID. We analyzed time and lineage between index infection and reinfection. We also analyzed the clinical outcome, severity of infection and vaccination status during reinfections.

**Findings:** In this study we confirmed that each wave of SARS-CoV-2 infections was characterized by a different viral variant showing a prevalence higher that 95%. We found that the fraction of reinfection is not linearly related to the average time of separation between waves with 40% of all the reinfections occurring at wave 5, the only wave with more than one SARS-CoV-2 variant with a prevalence higher than 80%. Regarding type of care 2.63% patients were considered ambulatory during the reinfection even when they were hospitalized during the index infection and only 0.78% presented the opposite behavior. Moreover, 6.74% reinfections transitioned from asymptomatic to mild or severe or from mild to severe; and 8.95% transitioned from severe to mild or asymptomatic or from mild to asymptomatic. The highest number of reinfections have occurred in unvaccinated patients (41.6%), followed shortly by vaccinated patients (31.9%). However, most reinfections occurred after wave 4 when the national vaccination efforts have reached 65% of the general population.

**Interpretation:** The analyzed data suggests a diminished severity of infection during reinfection either if transitions in disease severity or transitions in type of patient care are considered. Finally, we also observed an overrepresentation of unvaccinated patients in reinfections.

## Introduction

SARS-CoV-2 pandemia has lasted almost three years. Noteworthy, the number of infections continue to be important even when more than 68% of the global population has been vaccinated (https://ourworldindata.org/covid-vaccinations).

Some studies have characterized the reinfection rate over different periods of time showing varying degrees of reinfection rates, e.g, 0.2% between 1 June 2020 and 28 February 2021, 2.2% up to 1 year after previous infection, vaccine breakthrough infection rate of 5.6% with data cut-off on June 2021 and 6.7% vaccine breakthrough infection rate with a follow-up period of 277 days (1,2).

Reinfection risk has been shown to increase as the time to the first infection increases, reaching a maximum and stabilizing. Different time periods required to reach this stability have been reported from 277 days to 18 months at a maximum value of 6.7% and 18.86%, respectively (3,4).

Understanding disease severity of reinfections compared to primary infection is important to anticipate the burden of public systems and to aid on decision-making. Several reports have found that previous infection gives protection against severe reinfections and that risk of hospitalization and deaths diminishes in reinfections (5,6). A study found that the risk of having a severe reinfection was extremely low in persons previously infected compared to uninfected persons (1%)(5). Importantly, protection achieved by primary infection is comparable with that offered by vaccines (7).

However, the interpretation and application of all findings are complicated since virus variants have diverged over time and geographic location, vaccine distribution per vaccine type has been uneven around the globe, local vaccine administration strategies have sometimes been focused on specific population sectors, and vaccine efficiency per type is different. Besides, reports do not always categorize reinfection rate per vaccination status.

As the pandemia evolves reinfections cases are more common. In this study we analyzed the prevalence of SARS-CoV-2 variants over time in Mexico City during the infection waves. We also analyzed reinfection cases in Mexico City and the time between infections. We related the reinfection rate with the change in predominant SARS-CoV-2 variant. We also analyzed the clinical outcome, severity of infection and vaccination status during reinfections.

## Methods

### Data collection

This is a retrospective study analyzing data from the SISVER (respiratory disease epidemiological surveillance system) database, the mexican federal government central COVID-19 case reporting system. Briefly, this is a “line-list” case dataset, where each row corresponds to a single case. Only official public reporting institutions (either at the state or federal level) report to this system. As a member of the Federal Health System, our institution has access to the full database, including 130 variables for each case. For this work, we considered the following inclusion criteria: i) confirmed COVID-19 cases, having a positive result through either PCR or antigen testing, ii) records collected in medical units or testing sites located within the boundaries of Mexico City; and only considering cases with a reported residence in Mexico City and iii) cases collected up to 2022-07-16 (end of epidemiological week 2022-28). This amounts to 1,413,288 total cases analyzed.

This study was approved by the ethics and research committees of the Instituto Nacional de Medicina Genómica (CEI/1479/20 and CEI 2020/21).

### Identification of reinfections in the dataset

We define as a reinfection any secondary case with symptoms onset date at least 28 days later than the primary infection symptom onset date, based on the cutoff date used in (8).Tertiary and quaternary infections were also identified if they had symptoms onset date at least 28 days later than the previous infection. We used the Unique Key for Population Registry (CURP), which is a unique population identifier recorded in the dataset to uniquely identify each individual. Using this criteria, we found 88906 cases of people that had more than one confirmed infection (including the primary infections).

### Wave definition

The COVID-19 epidemic in Mexico, as in other parts of the world, has exhibited periods of high transmission or “waves.” The definition of such waves has been difficult due to changes in detection criteria. Therefore, we used a methodology first proposed in (9). Briefly, the authors looked at the (7-day rolling average) daily hospitalization counts, and considered a wave as beginning on the epidemiological week when the second derivative of this time series became positive, and ending on the epidemiological week when the second derivative of the time series became negative. We defined the periods between these waves as “interwave periods.” The start weeks for each wave and interwave periods are shown in Table 1.

**Table 1.** start weeks for each wave and interwave periods

| Start date (Epidemiological week) | Period |
| --- | --- |
| 2020-10 | Wave 1 |
| 2020-33 | Interwave period 1 |
| 2020-44 | Wave 2 |
| 2021-13 | Interwave period 2 |
| 2021-24 | Wave 3 |
| 2021-42 | Interwave period 3 |
| 2021-50 | Wave 4 |
| 2022-15 | Interwave period 4 |
| 2022-21 | Wave 5 |

### Variant prevalence analysis

For sequencing, we use Illumina and Oxford Nanopore platforms.

#### For *Illumina Sequencing*

The libraries were prepared using the Illumina COVID-seq kit, following the manufacturer’s instructions. First-strand synthesis was carried out with RNA samples, the synthesized cDNA was amplified using ARTIC primers V4, then was tagmented and adapted using IDT for the Illumina Nextera UD Indices Set A, B, C, D (384 indices) (Illumina, San Diego, CA, USA). Dual-indexed pair-end sequencing with a 150 bp read length was carried out on the NextSeq 2000 platform (Illumina, San Diego, CA, USA).

#### Oxford Nanopore Sequencing

Libraries were prepared according to ARTIC Midnight protocol PCR tiling of SARS-CoV-2 virus with rapid barcoding kit (SQK-RBK110.96) and sequenced on the GridION sequencing platform. We use the PCRT_9125_v110_revE_24Mar2021 protocol. 800 ng of DNA library was loaded into a primed R.9 flow cell (FLO-MIN106). MinKNOW software v.21.11.7 (Oxford Nanopore Technologies, Oxford, Oxfordshire, UK) was used to collect raw sequencing data and basecalling. Oxford-Nanopore Raw Data Processing and Sequencing Data Quality Assessment Basecalling and barcode demultiplexing were performed following the ARTIC protocol [https://github.com/artic-network/artic-ncov2019]. We regularly upload genomic and metadata information to GISAID (10).

### Clinical severity characterization

To assess whether post-primary infections are milder than the original infection, we used the symptom information contained in the SISVER database. The dataset has 20 binary variables to encode symptoms presented by the patient at the time of admission. Based on previous work (11) we labeled cases as severe if they reported any of the following symptoms: cyanosis, dyspnea, polypnea, or sudden symptom onset; or if the patient required intubation. Cases that presented no symptoms were labeled as asymptomatic. The rest of the cases were labeled as mild infections.

### Effects of vaccination on clinical presentation in reinfected patients

We performed a logistic regression model to predict the clinical severity of the second infection as follows :

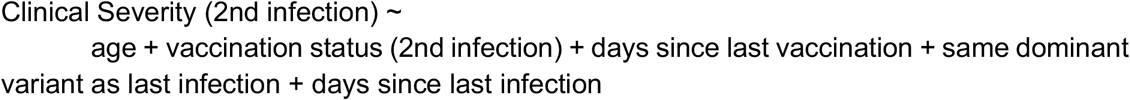

where the vaccination status were as follows: unvaccinated, if they had not received a vaccine previous to their infection; partially vaccinated, if they had received only one dose of a two-dose vaccine; vaccinated, if the infection occurred after receiving two doses; or boosted, if the patient had received an additional vaccine dose.

## RESULTS

### Each wave of SARS-CoV-2 infections was characterized by a different viral variant

First, we analyzed the prevalence of SARS-CoV-2 variants over time. The predominant SARS-CoV-2 variant has fluctuated across the different waves in Mexico City. Also, the duration and intensity, reflected by the number of cases, hospitalizations and deaths has varied.

During wave 1 (epidemiological week 2020-10 to 2020-32) B.1.1.222 was the predominant SARS-CoV-2 variant with a maximum incidence of 55.55%. B.1.1.222 was displaced by B.1.1.519 reaching a maximum incidence of 95.58% during wave 2 (epidemiological week 2020-44 to 2021-12) followed by the introduction of Delta which was the most prevalent in wave 3 reaching a maximum incidence of 100% (epidemiological week 2021-24 to 2021-41). After that, Omicron BA.1 reached a 100% prevalence during wave 4 (epidemiological week 2021-50 to 2022-14). And lastly during wave 5, Omicron BA.2 reached a maximum prevalence of 81.77% followed by the rapid introduction and spread of Omicron BA.5 which reached a 91.13% prevalence (epidemiological week 2022-21 - current week at the time of writing).

Number of cases reached a maximum during wave 4 (107,862 cases in week 2022-02) however the percentage of hospitalizations and deaths reached their maximum value at the start of the pandemia (wave 1) possibly due to a subestimation in the total number of cases followed by wave 2 with a maximum value of 12.29% hospitalizations and 8% deaths. Both indicators maintained a very low number at wave 5 with a maximum of 1.7% and 0.21%, respectively.

By other hand, the percentage of reinfections has been steadily increasing since 2021 epidemiological week 43, reaching a maximum of 9.78% by 2022 epidemiological week 28.

### Reinfections occurred mostly in the Delta and Omicron waves

We identify 42,963 reinfections throughout the pandemia. The general breakdown of the data is shown in Table 1. We calculated an average time between reinfection of 53 weeks. The first largest fraction of reinfection corresponds to 8,583 (20%) patients with index infections at wave 2 that were reinfected at wave 4, with a 59 weeks of time between reinfections. The second largest fraction of reinfections was observed in 5,889 (13.7%) patients from wave 2 who were reinfected in wave 5 with a 82 weeks of average separation between reinfections; and the third largest fraction of reinfections occurred in 5,880 (13.6%) patients who were infected for the first time in wave 4 and for the second time in wave 5 with a 23 weeks of average separation between infections (Figure 2 and Supplementary Figure 1).

**Figure 1.**
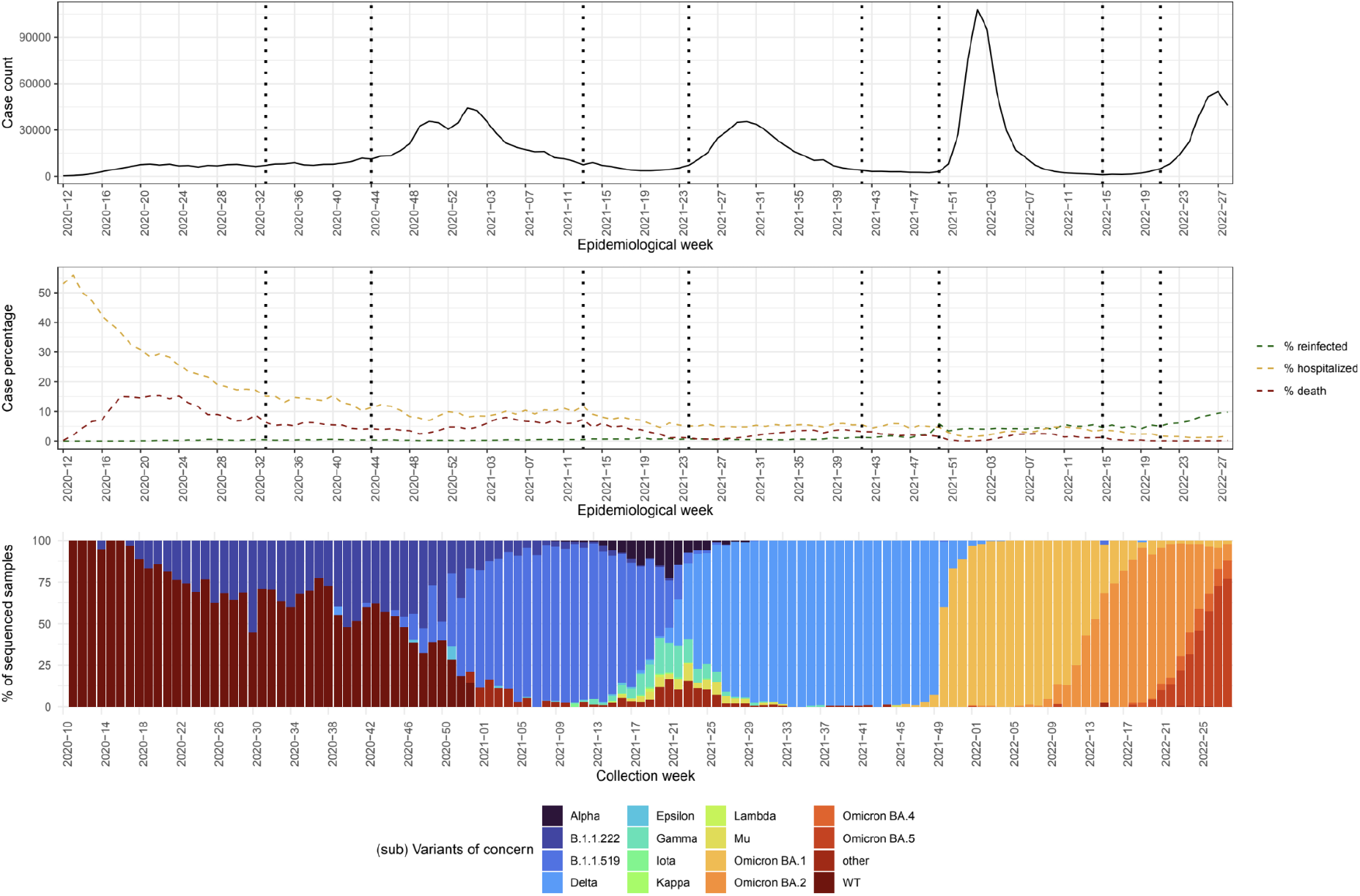
A) Number of cases per epidemiological week. b) Percentage of cases that belong to reinfections are shown in green. Also, percentage of cases that turned out in either death (red) or hospitalization (yellow) are shown. c) Percentage of sequenced samples per SARS-CoV-2 variant per epidemiological week.

**Figure 2:**
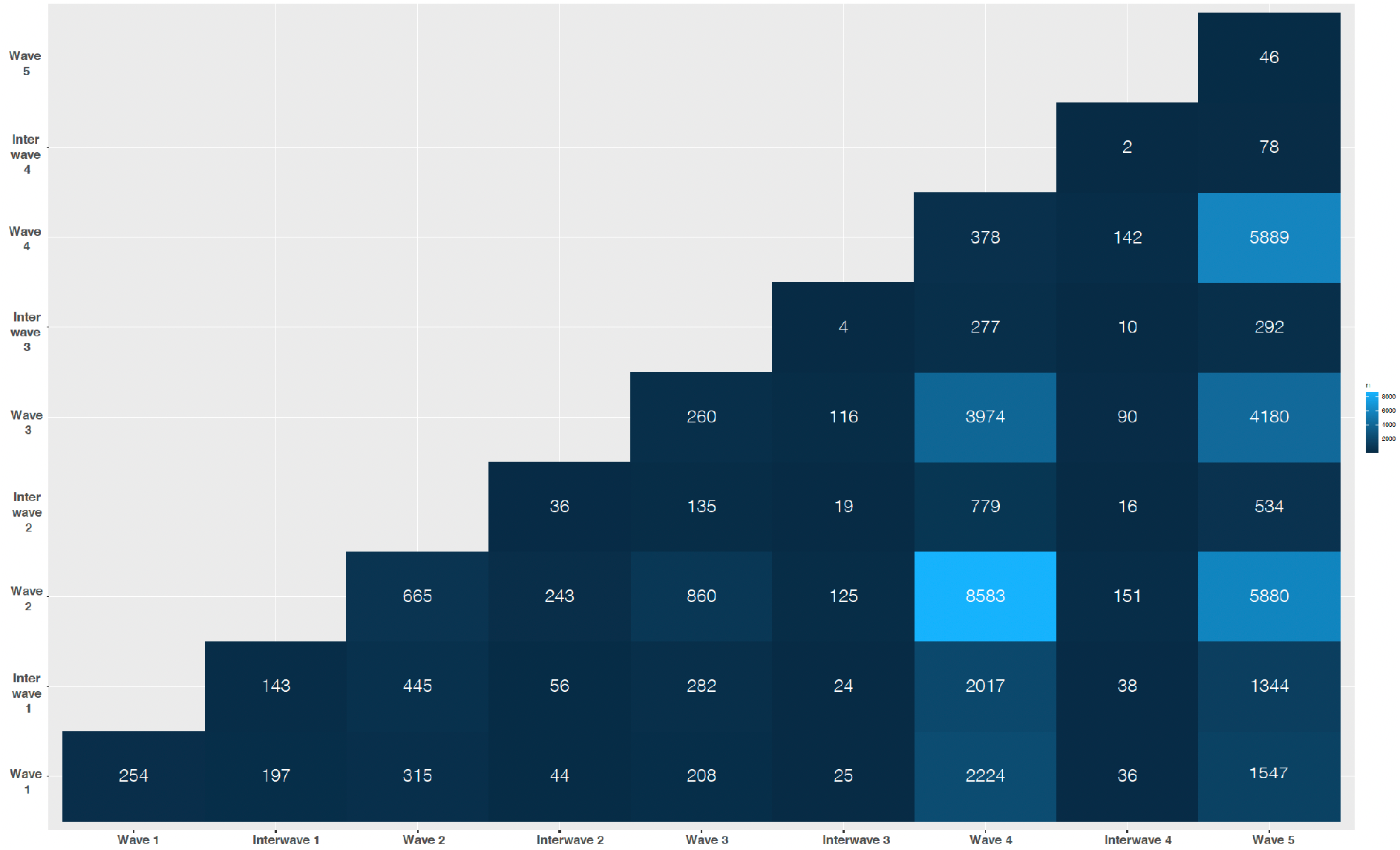
The number of reinfections across epidemic waves is shown. The x axis represent the time period of the index infection, the y axis represents the time period of the reinfection and the color of the tile indicates the number of reinfections in that specific combination of time periods.

So, the fraction of reinfection is not linearly related to the average time of separation between periods and seems to be occurring more often than expected at wave 5 of the pandemic accounting for 40% of all the reinfections. Wave 5 has been the only wave with more than one SARS-CoV-2 variant with a prevalence higher than 80% at different epidemiological weeks (Omicron BA.2 followed by Omicron BA.5).

### General data breakdown

Table 2 contains the basic breakdown of our reinfection dataset. The number of infections, the number of primary infections and the number of non-primary infection hospitalizations stratified by sex and age group.

**Table 2.** Breakdown of the reinfection dataset

|  | Male | Female | Total |
| --- | --- | --- | --- |
| Number of infections | 35347 | 53559 | 88906 |
| By age group |  |  |  |
| 0-9 | 582 | 506 | 1088 |
| 10-19 | 1798 | 2104 | 3902 |
| 20-29 | 8282 | 11469 | 19751 |
| 30-39 | 9709 | 14510 | 24219 |
| 40-49 | 7590 | 13249 | 20839 |
| 50-59 | 4957 | 8334 | 13291 |
| 60-69 | 1829 | 2665 | 4494 |
| 70+ | 600 | 722 | 1322 |
| Number of primary infections | 17482 | 26465 | 43947 |
| By age group |  |  |  |
| 0-9 | 309 | 271 | 580 |
| 10-19 | 957 | 1154 | 2111 |
| 20-29 | 4299 | 5931 | 10230 |
| 30-39 | 4742 | 7175 | 11917 |
| 40-49 | 3719 | 6482 | 10201 |
| 50-59 | 2359 | 3905 | 6264 |
| 60-69 | 815 | 1214 | 2029 |
| 70+ | 282 | 333 | 615 |
| Number of non-primary infection hospitalizations | 234 | 243 | 477 |
| By age group |  |  |  |
| 0-9 | 16 | 6 | 22 |
| 10-19 | 11 | 11 | 22 |
| 20-29 | 15 | 36 | 51 |
| 30-39 | 45 | 37 | 82 |
| 40-49 | 42 | 37 | 79 |
| 50-59 | 39 | 40 | 79 |
| 60-69 | 41 | 46 | 87 |
| 70+ | 25 | 30 | 55 |

### Clinical outcome and severity of infection during reinfections

We analyzed the clinical outcome, e.g. indicated by the type of patient care: ambulatory vs hospitalized, which is indicative of infection severity associated with reinfection. In Table 3 we present the number of reinfections per each clinical outcome, *e*.g. those that did not change clinical status during reinfection and those that transitioned from ambulatory to hospitalized and from hospitalized to ambulatory clinical outcome during reinfection. We found that in most cases 96.3% stayed as ambulatory in both infections. 2.63% patients were considered ambulatory during the reinfection even when they were hospitalized during the index infection. Only 0.78% presented and increased severity in its reinfection. And, 0.305% were hospitalized in both infections. This data suggests a diminished severity of infection during reinfection. However, this phenomenon cannot be solely associated with reinfection as most reinfections occur starting from wave 4 and vaccination started in wave 2 and has continued over time.

**Table 3.**
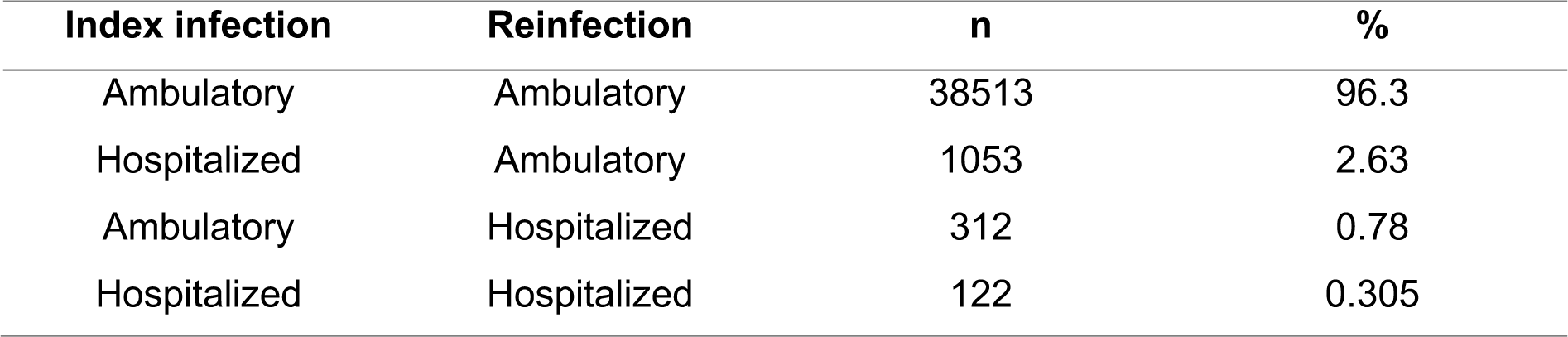
Transitions of type of patient care during reinfection

We also analyzed the severity of symptoms classified as asymptomatic, mild or severe during infection and reinfection (Table 4). Most SARS-CoV-2 patients present a mild infection regardless of the number of infections: 89.6% and 90.0% mild infections in the case of index infections and reinfections, respectively. Asymptomatic infections are not common in the federal SISVER database. However, this could be an underestimate given that most data in the database is derived from hospital and clinical centers that receive symptomatic patients. So, this number may not properly reflect the rate of asymptomatic infections in the general population. 84.34% reinfections did not change severity of symptoms during reinfection: 1.47%, 82.6, and 0.27% stayed as asymptomatic, mild or severe, respectively; 6.74% showed an increase in severity, ie. transitioned from asymptomatic to mild or severe or from mild to severe; and 8.95% showed a reduction in severity, ie. transitioned from severe to mild or asymptomatic or from mild to asymptomatic. So, reinfection tends to show a reduction in severity regardless of the wave in which the index infection or reinfection occurred (Supplementary Figure 2). This data is consistent with the diminished severity of infection during reinfection observed when the type of patient care is considered.

**Table 4.** Transitions of disease severity during reinfection

| Disease severity index infection | Disease severity reinfection | N | % |
| --- | --- | --- | --- |
| Asymptomatic | Asymptomatic | 631 | 1.47 |
| Asymptomatic | Mild | 2577 | 6.00 |
| Asymptomatic | Severe | 37 | 0.09 |
| Mild | Asymptomatic | 2728 | 6.35 |
| Mild | Mild | 35477 | 82.60 |
| Mild | Severe | 281 | 0.65 |
| Severe | Asymptomatic | 108 | 0.25 |
| Severe | Mild | 1008 | 2.35 |
| Severe | Severe | 116 | 0.27 |

**Tabla 5:** Regresion model covariates

| Covariate | Estimate | Std error | Statistic | Pvalue |
| --- | --- | --- | --- | --- |
| Status: fully vaccinated | 0.3333 | 0.1109 | -9.9075 | 0.0000 |
| status: partial vaccination | 0.3028 | 0.1407 | -8.4896 | 0.0000 |
| (Intercept) | 35.2291 | 0.4896 | 7.2753 | 0.0000 |
| days since last vaccination | 1.0018 | 0.0004 | 4.7688 | 0.0000 |
| Age | 0.9890 | 0.0024 | -4.5531 | 0.0000 |
| Dominant variant same as last infection | 0.5978 | 0.1791 | -2.8728 | 0.0041 |

Tabla 5: Regresion model covariates

### Vaccination status for reinfection

Given that later waves occurred during or after the governmental vaccination campaign, we decided to explore whether reinfections were occurring in vaccinated or unvaccinated individuals. The Mexican vaccination strategy uses a large array of vaccine products. Individuals could not choose the type of vaccine that they would get, but were assigned one based on their age group and place of residence. So, the data was not stratified per vaccine brand to avoid any unintended grouping by age group or place of residence. Due to this complexity, we decided to label cases as unvaccinated, if they had not received a vaccine previous to their infection; partially vaccinated, if they had received only one dose of a two-dose vaccine; vaccinated, if the infection occurred after receiving two doses; or boosted, if the patient had received an additional vaccine dose.

Most index infections occurred at unvaccinated patients (35,399 out of 40,120) which is expected given that most index infections occurred at wave 2 before the national vaccination efforts reached a significant fraction of the population. From these infections, the highest number of reinfections have occurred in unvaccinated patients (41.6%), followed shortly by vaccinated patients (31.9%). However, most reinfections occurred after wave 4 when the national vaccination efforts have reached 65% of the general population (ourworldindata.com) which is an underestimation of the progress of the vaccination program in Mexico City. This implies an overrepresentation of unvaccinated patients in reinfections. Interestingly, only 14.8% percent of reinfections occurred in patients with a boost dose of the vaccine.

**Figure 2:**
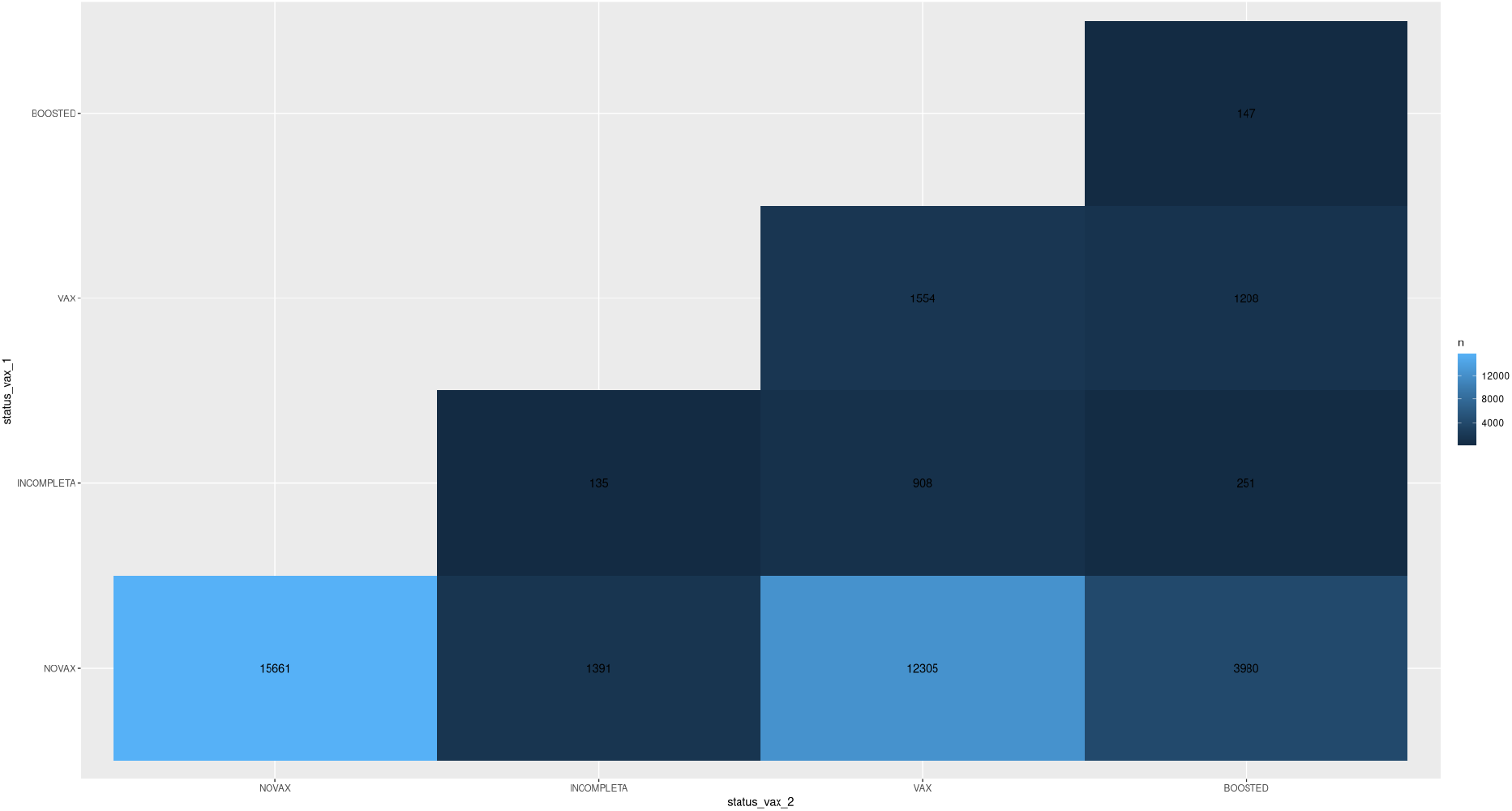
The number of reinfections per each combination of vaccination status. The x axis represent the vaccination status of the index infection, the y axis represents the vaccination status of the reinfection and the color of the tile is proportional to the number of reinfections for that specific combination

### Regression model to predict severity of disease

We also train a regression model to predict severity of disease during reinfection taking as covariates vaccination status, date of last vaccination, age, and the SARS-CoV-2 variant (only omicron BA.2 and omicron BA.1 were considered as covariates) (Table 3). We found that the vaccination status, the days since last vaccinations and the age of the patient were the covariates statistically related to severity of disease during reinfection

## DISCUSSION

For some viruses, the first infection generates lifelong immunity preventing reinfections, for seasonal coronaviruses this immunity is shorter, generating reinfections within the first 6 to 12 months (12). The ability of SARS-CoV-2 to mutate and generate new variants has led to reinfections worldwide. The first case of reinfection occurred in August 2020 (13), and the number of cases will continue to increase as new variants are generated that evade the immune response previously generated by another variant.

This paper is the first, and largest, to cover cases of COVID-19 reinfection in Mexico City, raising questions regarding vaccination and exploring the clinical outcome of patients.

In Mexico City, we identified five waves of infection caused by five different variants. The first and second waves were caused by B.1.1.222 and B.1.1.519, variants that only circulated with higher prevalence in Mexico, while in other parts of the world there was a diversification of variants. Waves 3, 4 and 5 caused by Delta, BA.1 and BA.5 respectively also caused waves of infections around the world (14).

The infection rate was 2.43% (44,959 reinfections of 1,847,662 positive cases). In comparison with other studies, this infection rate is higher. Of the 42,963 reinfections detected, 33.7% occurred in Omicron BA.1 and BA.5 waves of infection, its variants already recognized for the gain of amino acid substitutions in spike protein (L452Q, L452R and F486V) associated with evasion of immune response and high affinity to ACE2 receptor (15,16). The average number of days between primary and secondary infection in patients in Mexico City was 53 weeks, the period reported in a meta-analysis of 577 reinfection cases in 22 countries was 63 weeks (17).

Although reinfections represent a low percentage of newly diagnosed cases, they will become increasingly frequent given the generation of new variants that evade the immune response previously generated by vaccination or infection such as BA.2.75, BQ.1, BQ.1.1 and XBB, variants newly identified in November 2022 and already circulating worldwide with positive *in vitro* assays for evasion of antibody-mediated immune response (18–20).

Only 0.78% of outpatients required hospitalization for their second infection; we have no further information about the diagnosis of hospitalized patients and/or the symptoms they presented. In a study of reinfections it was observed that 63 patients in their second infection presented: pneumonia and acute renal damage(21), while in another study 48 patients debuted with respiratory failure, thromboembolism and sepsis (22).

It is known that the accumulation of SARS-CoV-2 infections results in a hazard ratio of hospitalization and death of 3.32 and 2.17, respectively (23). Accumulation of sequelae in multiple organs (lung, cardiovascular system, gastrointestinal system, kidneys, musculoskeletal and neurological), as well as mental health damage have been reported (24). Given that SARS-CoV-2 will continue to mutate for years to come, new variants with the potential to generate reinfections will continue to emerge, which could lead to a greater burden on the public health system given the comorbidities that occur in patients with reinfection, and its prevention will be one of the greatest future challenges in public health.

## Data Availability

All data produced of SARS-CoV-2 cases sequenced by INMEGEN are deposited in GISAID.

https://gisaid.org

## DATA AVAILABILITY

All data produced of SARS-CoV-2 cases sequenced by INMEGEN are deposited in GISAID.

## AUTHOR CONTRIBUTIONS

Conceptualization and study design: GAJ, LGR, ACT. Data acquisition: GAJ, LGR, SC, ACR, JAF. Data analysis/interpretation: GAJ, LGR, ACT. Statistical analysis: GAJ, LGR. Manuscript drafting: GAJ, LGR, ACT. Manuscript Review & Editing: AHM, LAH. Funding Acquisition: ACT, AHM, LAH.

All authors have read and agreed to the published version of the manuscript.

