## Supplementary Figures for "COVID-19 Reinfections in Mexico City: Implications for public health response"

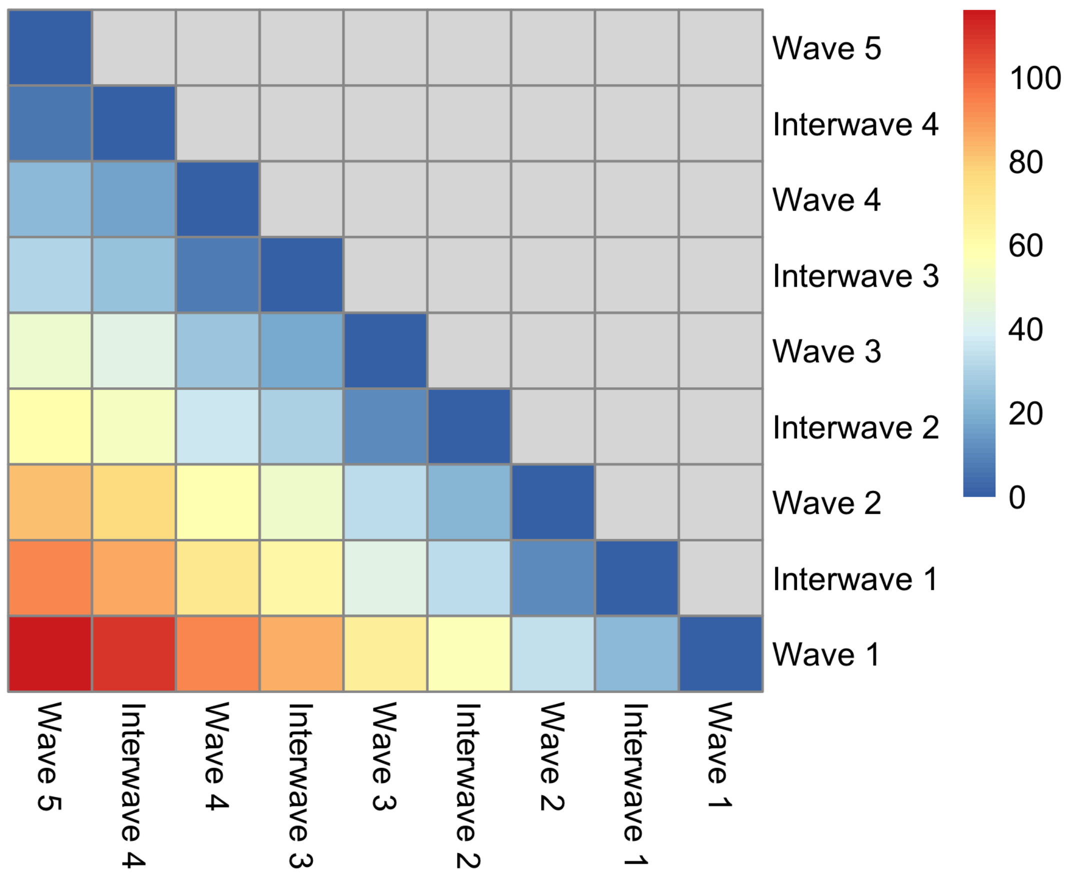


Supplementary Figure 1. Time between waves

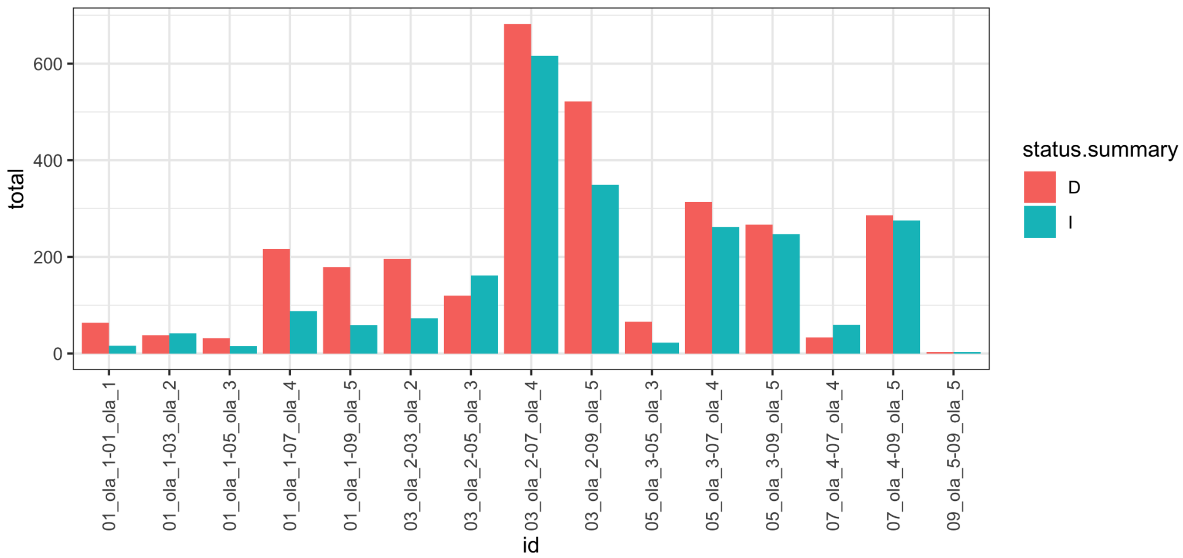


Supplementary Figure 2.  Reinfection tend to show a reduction in severity
